# Requirements for the delivery of an Australian Rapid Access Chest Pain Clinic

**DOI:** 10.1101/2024.07.31.24311327

**Authors:** Annika Wilson, Laura Sutton, Rose Nash, J. Andrew Black, Senali Jayasinghe, James E. Sharman, Niamh Chapman

## Abstract

**Background:** Rapid Access Chest Pain Clinics (RACPCs) are outpatient cardiac services designed to promptly assess and manage patients experiencing chest pain. Despite the establishment of 25 RACPCs across Australia, a standardised implementation framework has yet to be developed. This study aimed to identify the core components of successful delivery of an existing RACPC.

**Methods:** A qualitative process evaluation study was conducted at a RACPC in a metropolitan, tertiary hospital in Tasmania, Australia, from November 2022 to July 2023. Clinical observations and semi-structured interviews were conducted with seven RACPC clinicians. Deductive data analysis was undertaken according to a Context-Mechanism-Outcome framework.

**Results:** Core components of successful RACPC delivery included: (1) a multidisciplinary team-based approach to care with discreet clinical roles; (2) timely patient review by RACPC clinicians within 30 days of referral; (3) embedded patient education; (4) ongoing clinical team training and education; and (5) a shared understanding of the RACPC service’s identity and purpose. Challenges to RACPC delivery were also identified and included resource constraints and administrative burdens.

**Conclusions:** Successful delivery of a RACPC model of care relies on a range of interrelated factors. These findings align with the broader theme of ongoing health service evaluation as a driver for continuous quality improvement and care standards within RACPCs. Further research aimed at developing and implementing effective strategies to enhance service delivery is needed to determine a national model of care.

## Introduction

Chest pain is the second most common presentation to Emergency Departments (ED) across Australia, accounting for 6% of hospital admissions, but with only 15% of these cases being diagnosed as acute coronary syndrome [1,2]. Similar findings have been reported in the USA [3] and the UK [4], contributing to a global issue of ED overcrowding [5]. Australian guidelines recommend prompt assessment and management of patients presenting to ED with low-risk chest pain in outpatient settings [6]. Rapid Access Chest Pain Clinics (RACPCs) have emerged as an effective model to ensuring safe discharge and timely follow-up for these patients. Early experiences indicate a low risk of cardiovascular events within 30 days, supporting the outpatient approach [7–10]. Despite the presence of 25 RACPCs across Australia, the absence of a standardised care framework for their implementation has led to potential variability in service delivery [11].

There remains a dearth of understanding about RACPC operational mechanisms, implementation processes, and the contextual factors influencing outcomes. The limited qualitative evaluations available have primarily focused on the perceived value of RACPCs among referring providers [12], as well as patient outcomes and experiences [13]. Health service evaluation is an important process to identify the critical components that facilitate safe and sustainable delivery of care, particularly for complex multi-modal health interventions such as RACPCs [14,15]. Realist evaluation has emerged as an effective methodology in health services research for assessing complex interventions, aiming to answer the questions: ‘what works for whom, in what context, to what extent, how, and why’ [14]. To our knowledge, no studies have yet been published that evaluate the contextual factors and mechanisms affecting RACPC outcomes.

This study aimed to conduct a realist process evaluation of an existing RACPC from the perspective of its clinical staff. By doing so, this study seeks to explore the following research question: What are the core components for successful delivery of a RACPC?

## Materials and Methods

### Study setting and study design

A single site evaluation was undertaken from a RACPC at a metropolitan, tertiary hospital in Tasmania, Australia. The RACPC was established at the Royal Hobart Hospital in 2014 where the model of care has been described previously [7,11]. The RACPC involves a multidisciplinary clinical team comprised of a clinical nurse (n=1), senior cardiologist consultant (n=1), and a rotating medical registrar and resident (n=2). Observations of RACPC service delivery were undertaken according to a pre-determined checklist to develop assumptions regarding the contextual and mechanistic factors that impact on RACP delivery and its outcomes. Subsequently, semi-structured interviews were undertaken with RACPC clinical staff to explore their perceptions of the contextual factors and supporting mechanisms identified during the observations. We followed the realist heuristic where implementation factors were used to map the relationship between contextual conditions and mechanistic factors that lead to intended or unintended outcomes of RACPC delivery [14]. In this study, mechanisms have been delineated as the imperceptible, implicit processes within individuals’ minds concerning the intervention, and encompasses their perceptions of how and why the intervention works [16]. Consistent with realist evaluation, a pragmatist worldview was adopted by the researchers, meaning the research was problem-centred, real world and solutions orientated. As such realist evaluation was the most appropriate method to answer the research aims and acknowledges that the relationships between the context, mechanisms and outcomes (CMOs) in a RACPC setting be explored.

### Sample and recruitment

A purposive sampling strategy was employed to recruit key clinical staff members involved in the delivery of the RACPC at the Royal Hobart Hospital between November 2022 to July 2023. All clinical staff were contacted via email or approached in-person by a member of the research team. During the study period, staffing changes and leave coverage made two additional staff members (a medical registrar and nurse) eligible to participant. The total sample recruited was seven. Written informed consent was obtained from all participants.

### Clinical observations

Using the previously described model of care for RACPC delivery in this setting [7], an observation checklist was developed to capture the clinical environment related to patient referral, nurse assessment, doctor consultation, follow-up investigations, referrals, and clinical note taking (see Appendix A). Direct observations of clinical workflow were carried out by three separate investigators (LS/SJ/AW) and involved multiple observations to capture variations in patient management.

### Interview procedure

A semi-structured interview schedule was developed based on the previously described model of care, a literature scan, initial observations, and the CMO framework (Appendix B). The interview schedule was conducted with key RACPC staff and included: (1) the level of services offered to patients; (2) the efficiency and effectiveness of the service processes; (3) the management, resourcing, and communication with the service; and (4) any positive or negative attributes encountered.

Interviews were audio-recorded and transcribed verbatim by a single author (AW). Based on initial analysis of the seven interviews, two follow-up interviews were conducted with a senior consultant cardiologist and a medical registrar to further explore and/or clarify themes. Interviews ranged between approximately 14 to 28 mins, with a total duration of 3 hrs 05 mins.

### Researcher reflexivity

The study team comprised experts in cardiovascular research (NC/JS/JAB), health service implementation (NC/JAB/JS/RN/LS), clinical practice (JAB/SJ), and qualitative research methods (NC/JS/LS/RN/AW). Researcher reflexivity was addressed by disclosing background information about investigators to participants before the interviews.

### Data analysis

Data analysis was undertaken iteratively throughout data collection using a deductive thematic approach [17] and framework analysis [14]. A codebook was pre-determined by the authors using *a priori* codes based on an extensive review of the relevant literature according to CMO domains, which underwent further development following the initial coding of three transcripts and two observations (Appendix C). All interview transcripts were coded and analysed by a single author (AW) in NVivo (version 12). Six transcripts were selected at random and coded by two authors (NC/LS) for quality assurance. Any discrepancies were discussed between authors (NC/LS/AW) until consensus was reached. The analysis team met regularly to discuss the key themes and findings.

Preliminary findings were shared during a Royal Hobart Hospital cardiology team meeting, involving both participant and non-participant staff to incorporate a wider clinical perspective in the data interpretation. Feedback from the meeting prompted further modifications to data analysis and interpretation and contributed to a better understanding of the RACPC process.

Study procedures were undertaken in accordance with local ethical approval and site-specific governance (Ref: H0018103) and reported according to the RAMESES II guidelines on the reporting of realist evaluation research [18].

## Results

Nine interviews (involving seven interviews and two follow-up interviews) and 24 clinical observations were conducted across the study period and involved seven participants: a senior consultant cardiologist (n=1), medical registrars (n=3), medical resident (n=1), and nurses (n=2).

Framework analysis led to the identification of five factors to support positive RACPC delivery: (1) multidisciplinary team-based approach to care; (2) timely patient review; (3) embedded patient education; (4) clinical team education and training; and (5) shared understanding of RACPC service identity and purpose. An additional CMO factor was identified and described aspects that could negatively impact RACPC delivery: (6) Lack of organisational support and resources. Themes and their corresponding CMO factors are presented in Table 1 and described in more detail below.

**Table 1.**
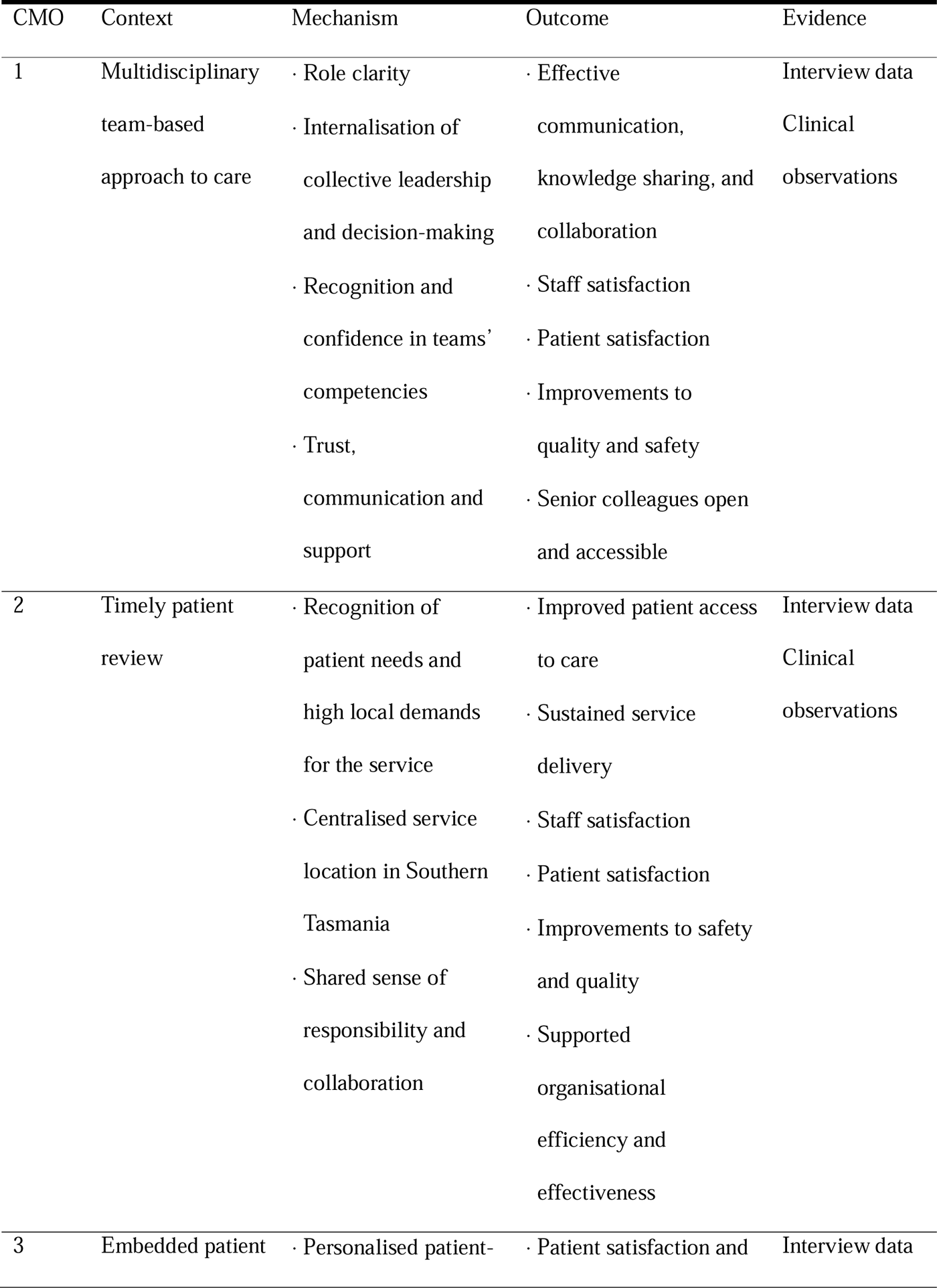
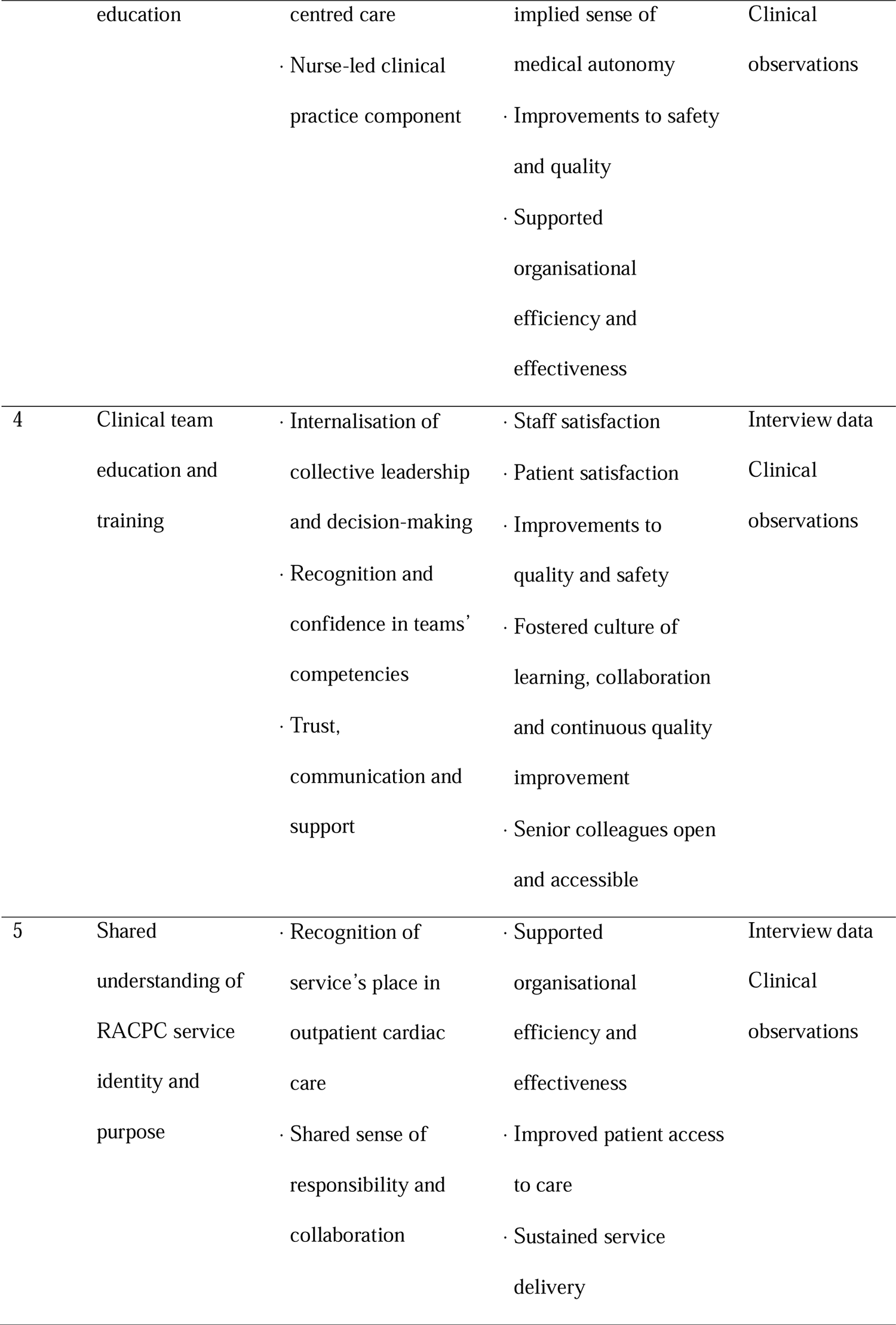
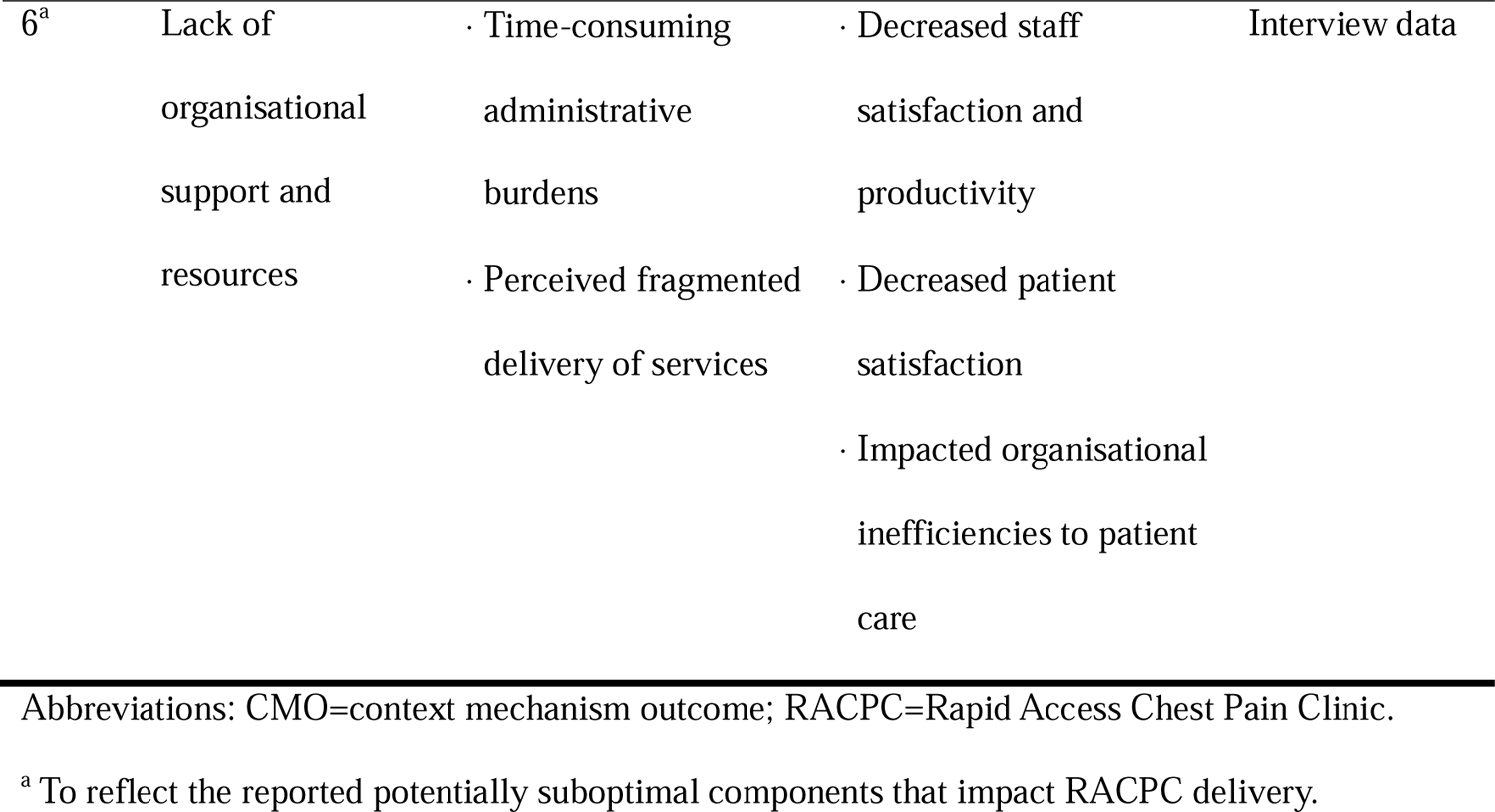
Potential CMO factors from in-depth interviews with RACPC clinicians.

### 1: Multidisciplinary team-based approach to care

Adopting a multidisciplinary team-based approach to care delivery was a key determinant of successful delivery of the RACPC. This approach is rooted in the recognition that addressing complex health issues, such as cardiac care, necessitates the expertise and insights of diverse healthcare professionals working together. Several key mechanisms emerged and included clear delineation of roles, recognition of collective leadership and shared decision-making, and confidence in the competencies of all RACPC clinicians. In the context of the RACPC, the significance of these mechanisms became more evident: the RACPC is staffed by a dedicated nurse (n=1) who coordinates the clinic (including patient administration and communication) and provides an initial absolute cardiovascular risk assessment and patient counselling for smoking cessation, weight loss, and exercise as required. Residents and registrars (n=1-2 per week) provide clinical assessment of patients and facilitates investigative strategy (Fig. 1). A senior consultant cardiologist (n=1) leads the team, offering specialised expertise and oversees the diagnosis, treatment planning and follow-up of patients. This approach was clearly described by Interviewee 7, who said: *“We all work as a team. It’s a collective approach so we have to make sure we’re all on the same page.”*

**Fig 1.**
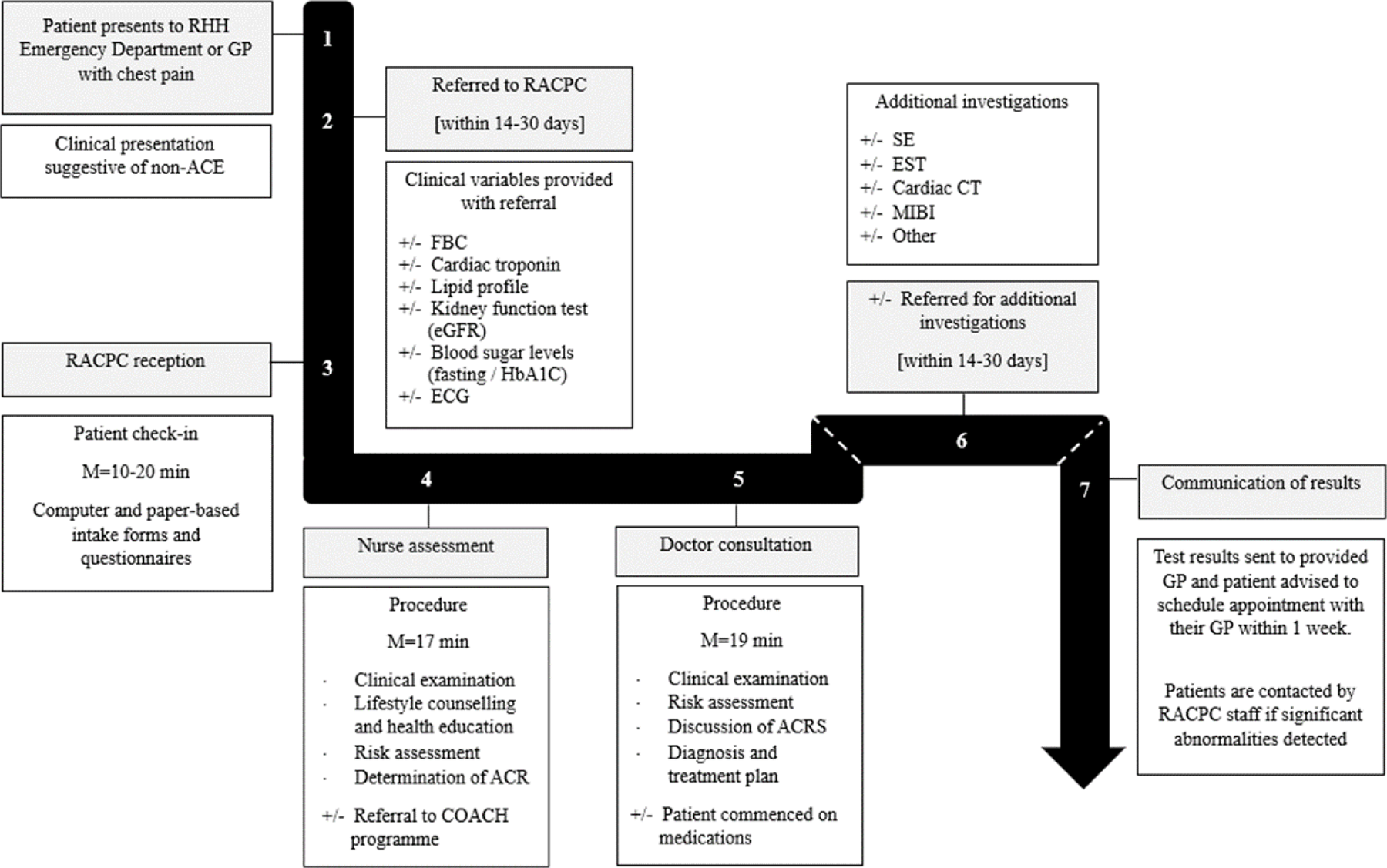
The RACPC model of care. Abbreviations: ACR=Absolute cardiovascular risk; ACRS=Absolute cardiovascular risk score; CT=Computerised tomography; ECG=Electrocardiogram; eGRF=Estimated globular filtration rate; EST=Exercise stress test; FBC=Full blood count; GP=General practitioner; M=Mean; MIBI=Myocardial perfusion scan; Non-ACE=Non-Acute Coronary Event; RACPC=Rapid Access Chest Pain Clinic; RHH=Royal Hobart Hospital; SE=Stress echocardiogram.

The observed outcomes in line with this multidisciplinary approach included improved communication, knowledge sharing and collaboration, alongside higher levels of staff and patient satisfaction, enhanced quality and safety, and increased accessibility to senior colleagues. Interviewee 3 shared their experience:

> *In terms of human resourcing, at a team-based level, it’s good that we’ve got [Senior Consultant]. I think [Senior Consultant] the right person for it cause not only is he the standout lead and our vehicle liaison person for like for the state, you know he’s an interventionist and you can oversee all of these referrals. So he’s like kind of the right person for leading the service. Next, we’ve got a nurse who’s dedicated to RACPC, as the patient load is humongous. So that’s really good. And then having dedicated time slots where a few doctors come and can just smash it out and having a clinic that services one issue… I find it useful.*

### 2: Timely patient review

Timely patient review was viewed as critical within the RACPC, driven by the mechanisms of high patient needs and demands for the service, coupled with a shared sense of responsibility and collaboration among clinical staff. In context, operating the RACPC for at least two days per week was deemed essential to manage the influx of referrals from the ED and broader community providers (such as referring GPs), and ensured that patients were more likely to receive timely and safe follow-up care. These findings were reinforced via the clinical observations which recognised that the RACPC’s centralised location and co-location to the Royal Hobart Hospital was an important factor in providing accessible care to patients across the greater Hobart and southern Tasmania region.

During the interviews, clinicians also stressed the necessity of reviewing patients within 30 days to align with the expectations of a rapid access system, facilitating early intervention and mitigating potential complications or treatment delays. The outcomes observed in these contexts were twofold: timely patient review not only facilitated prompt evaluation of cardiac concerns but also addressed non-cardiovascular symptoms, thereby potentially decreasing the likelihood of unnecessary investigations and overservicing, as Interviewee 2 explained:

> *I think the speed at which patients are seen, I think that can then take a lot of burden off both primary healthcare, but also the general cardiology clinics. And then knowing that there’s going to be good follow up of these people and the results of their investigations and hopefully - I would hope - preventing adverse outcomes as a result of that*.

Overall, the outcomes associated with timely patient review encompassed improved patient access to care, sustained service delivery, heightened staff and patient satisfaction, enhanced safety and quality measures, and supported organisational efficiency and effectiveness. These outcomes, and what the RACPC offered, were explored in the following quotes:

> *I think definitely for the RACPC it allows patients to be seen much quicker upon discharge. I think the waiting list for many of the other clinics is very long and a lot of patients also struggle to access their GP. So I think that I think it’s an important service to ensure that patients are seen in a in a timely fashion. – Interviewee 6*.

And,

*Timely access to consultation with clinicians in outpatient setting, timely access to cardiac investigations to rule out the cause of the chest pain. And probably a more evidence-based approach to interpretation of those investigations… And adequate or probably better communication with GPs than exists at hospitals. – Interviewee 4*.

### 3: Embedded patient education

The pivotal role of embedded patient education was raised as a crucial component of the RACPC service and viewed as an opportunity to implement strategies for primary and secondary prevention of cardiovascular diseases. Findings from participant interviews and clinical observations revealed two main opportunities for patient education: before and during the initial appointment at the RACPC (see Fig 1). Providing education prior to the presentation through information leaflets and referring provider discussion was thought to equip patients with essential knowledge of the service and was intended to prepare them for their upcoming consultation. Delivering education during the appointment via education materials and clinical consultation ensured patients were informed of their condition, diagnostic procedures, absolute cardiovascular risk, and treatment options, which was thought to empower them to actively participate in their care. These integrated efforts are mechanistically underpinned by principles of personalised, patient-centred care and nurse-led clinical practices. The outcomes of this approach are manifold, and included heightened patient satisfaction, improvements in safety and quality metrics within the clinic, and streamlined workflow processes. As Interviewee 5 said:

> *I’ve always been a big advocate for lifestyle advice and giving patients the tools to make those decisions themselves, and this job allows me to do that*.

### 4: Clinical team education and training

Despite no formalised training programme currently implemented within the RACPC, clinical staff described positive experiences related to clinical team education and training as a component of their time working in the RACPC. Within the RACPC context, these experiences seemed to reflect the unique team and leadership dynamics, and close association with the cardiology clinics at the Royal Hobart Hospital. For example, a more junior staff member reported feeling more supported and equipped to handle the complexities of cardiac care due to an ability to openly communicate and seek advice from other clinical staff members, particularly the senior consultant cardiologist. As Interviewee 2 described:

> *It’s good having a consultant here at every clinic who, as a more junior staff member, I can then discuss cases with if I feel like I’m uncertain or just want a bit of reassurance*.

Additionally, departmental meetings involving co-located cardiac clinics are held on a weekly basis and were reported to include opportunities to confer on cases, discuss updates on cardiac research and guidelines, and reinforce policies and protocols. Mechanisms supporting this finding included a sense of collective leadership and decision-making, trust and communication, and a recognition and confidence in teams’ competencies. Notable outcomes of this approach were the enhancement of staff and patient satisfaction levels, improvements to quality and safety, and fostered culture of continuous learning and improvement that supported adaptive and consistent cardiac care. To illustrate:

> *The important thing is that they [clinical staff] are contributing to an important part of their training and addresses a key issue with the accreditation of junior staff training, which is that they don’t have enough clinic exposure… So it [RACPC] is useful for that - it’s good for training. It’s also good for medical student teaching as well. – Interviewee 1*.

### 5: Shared understanding of RACPC service identity and purpose

The importance of a shared understanding of RACPC service identity and purpose was viewed as a unique feature of this model of care by participants. Key mechanisms contributing to this context include the recognition of the service’s place in outpatient cardiac care, as well as the cultivation of a shared sense of responsibility and collaboration among clinical staff. Within the RACPC, clinical staff have embraced and celebrated its distinct identity within outpatient cardiac care. They acknowledged the unique values and processes inherent to the RACPC, and recognised its role in delivering high-quality, preventive, and patient-centred care. Moreover, staff members appeared to hold the RACPC in high regard for its reputation as a forerunner in implementing innovative and efficient healthcare practices and service models within the Tasmanian Health Service, with Interviewee 2 explaining:

> *RACPC seems to know its role, and like what the clinic is here to do, seems to be pretty clearly defined. So we’re able to stick to that area and therefore see people efficiently. – Interviewee 2*.

As a result, the outcomes of this shared understanding encompass a supported organisational efficiency and effectiveness through the decongestion of wait times, ED overburden and GP follow up, and a sustained service delivery over time by ensuring patients have improved access to timely and comprehensive care. As interviewee 2 described:

> *I think from the perspective of an emergency department clinician, it is reassuring to know that there is definite follow up booked for these patients and that [the RACPC] is reasonably quickly accessible. So I think that’s definitely a good thing and probably reduces the load or burden on GP follow up. – Interviewee 2*.

### 6: Lack of organisational support and resources

Lastly, challenges arising from a lack of organisational support and resources within the health services were highlighted as risks to RACPC delivery. Mechanisms contributing to this context include burdensome administrative tasks and a perceived fragmented service delivery model. To illustrate, challenges with ordering additional investigations and time-consuming administrative tasks were frequently reported. Several participants highlighted challenges with the process of the request, execution, and receipt of results for additional investigations. For context, referral processes are not standardised between departments and external diagnostic centres. Use of paper referrals at the time of the study period (now superseded by e-referral processes) were reported to make tracking referral pathways difficult, and follow-up cumbersome. Senior staff reported that a large amount of their clinical time in the RACPC was consumed by these time-intensive administrative tasks, including note taking and documentation of the consultation, referral coordination, and following up of test results where much of the workload fell to the clinical nurse and senior consultant. This administrative load impacts on capacity for patient care, resulting in diminished job satisfaction and reports of heightened stress levels. For example:

> *I would say it’s like a hamster wheel. It’s got all these different things. So, the forward planning, the retrospective, and then the current…Yes, I mean it is just constant, constant, attention to past, present and future patients. – Interviewee 5*.

Additionally, the capacity of the clinic was limited by the availability of consultation rooms. The clinic was usually overstaffed by at least one clinician compared to the number of consultation rooms available. Availability of space within the whole health service was limited and scheduling conflicts with co-located clinics were reported by the RACPC clinicians. Overall, this limitation was reported to impede service efficiency by potentially resulting in treatment delays or reduced accessibility to care. Interviewee 1 described their experience as follows:

> *So, space is limited. I mean we’re essentially in direct competition with other clinical services for space, so it’s been a lot of effort to claw into the space that we’ve got; and that can basically be very quickly reallocated or taken away. And then obviously there’s a critical number of doctors and clinic rooms we need to be able to function as we have to be able to see people within a timely manner. – Interviewee 1*.

## Discussion

This study describes the requirements for the successful delivery of a RACPC model of care and marks the first to conduct a realist process evaluation through the perspectives of its clinical staff. The six interrelated factors identified in this research provide a quality checklist for the other 25 RACPCs in operation across Australia and inform the future development of a standard of care guidelines to support future RACPC service implementation to ensure consistent service delivery.

Multidisciplinary team-based care was critical to achieving comprehensive and efficient cardiac care within the RACPC, with integrated clinical education and training emerging as a contextual enabler to supporting these outcomes. In healthcare, professions are typically trained in silos, often lacking training on how to effectively collaborate within multidisciplinary teams [19]. National guidelines recommend the implementation of multidisciplinary team approaches to enhance guideline adherence, support informed decision-making, and facilitate patient-centred care [6]. Within this study, the clinic’s dedicated staff collaboratively contributed to a well-coordinated and efficient workflow and fostered a culture of continuous learning and quality improvement. The transparent delegation of responsibility and shared decision-making, facilitated by discreet clinical roles, ensured clear communication and coordination of tasks utilising an enhanced collaborative approach. This aligns with existing research that highlights the effectiveness of team-based care in healthcare settings including patient satisfaction and enhanced clinical outcomes [20,21].

A clear service identity within the RACPC and timely patient review served as distinctive mechanisms for achieving RACPC outcomes. The RACPC’s strong identity, recognised by clinical staff for its values, efficient practices, and leadership as an innovative and evidence-based healthcare model, contributed to a shared sense of responsibility and purpose among the team. Notably, healthcare settings actively engaged in research often report improved patient outcomes, reinforcing the idea that the RACPC’s emphasis on evidence-based practices and continual quality improvement plays a crucial role in its success [22]. The focus on timely patient review in the RACPC, operating a minimum of two days per week, further underscored a commitment to maintaining a prompt and safe follow-up period. This finding aligns with the expectations of an established time threshold for patient review within 14-28 days within national guidelines [6] supporting the outpatient approach. However, in practice, review times were often extended as patients preferred convenience over promptness. This trend is consistent with observations in other RACPCs [7,10] and is considered safe.

Our study highlights the integral role of embedded patient education within the RACPC, aligning with broader strategies for primary and secondary prevention of cardiovascular diseases. Informed patients are more likely to adhere to prescribed treatment plans, adopt lifestyle modifications, and actively engage in preventative measures [23]. This proactive involvement is crucial in the context of cardiovascular health, where patient lifestyle choices play a significant role in long-term outcomes [24]. The integration of patient and clinician education within the RACPC also aligns with broader healthcare quality standards including a person-centred approach, clear organisational purpose, and leadership in healthcare delivery [25].

## Limitations

The study had several limitations. While all eligible clinical staff involved in RACPC delivery were recruited, the purposive sampling method may introduce selection and social desirability biases, limiting the representativeness of perspectives within the broader healthcare community. Future service evaluations should include interviews with referring providers and patients. Additionally, the findings may be specific to the RACPC model studied, so caution is needed when applying these results to different settings. To enhance the generalisability of the findings, future research should explore diverse healthcare contexts and include larger, more representative samples.

## Conclusions

This study demonstrated that the successful delivery of a RACPC model of care relies on several interrelated factors. As the first exploration of its kind, it provides invaluable insights into RACPC delivery from the perspectives of clinical staff at an Australian hospital. The findings underscore the importance of ongoing health service evaluation to maintain and improve patient care quality. Further research is needed to develop and implement effective strategies to overcome barriers and enhance service delivery.

## Data Availability

All data produced in the present study are available upon reasonable request to the authors

## Acknowledgements

We thank the participants and staff of the Royal Hobart Hospital Cardiology Department for their valuable input.

## Declaration of funding

This study is supported by a project grant from the Royal Hobart Hospital Research Foundation [22–202].

## Conflicts of interest

The fourth author (JAB) is an employee of the Royal Hobart Hospital Cardiology Department but was not involved in the analysis of the study. The remaining authors have no conflicts of interest to declare.

## Notes

### Competing Interest Statement

The fourth author (Dr J. Andrew Black) is an employee of the Royal Hobart Hospital Cardiology Department but was not involved in the analysis of the study. The remaining authors have no conflicts of interest to declare.

### Funding Statement

This study is funded by a project grant from the Royal Hobart Hospital Research Foundation [22-202]

### Author Declarations

Human Research Ethics Committee of the University of Tasmania gave ethical approval for this work (Reference number: H0018103).

