## Appendices for "Requirements for the delivery of an Australian Rapid Access Chest Pain Clinic"

**APPENDIX A. DATA COLLECTION CHECKLIST (CLINIC OBSERVATIONS)**

Date:	Observer:	Patient no:

**Step 0: Referral**

1. Patient arrival time:
2. Source: GP / ED
3. Clinical variables provided with referral:

- ☐ Full Blood Count
- ☐ Cardiac troponin
- ☐ Lipid profile
- ☐ Kidney Function test (eGFR)
- ☐ Blood sugar levels
- Fasting
- HbA1C
- ☐ ECG

4. Comments

**Step 1: Nurse Assessment**	**Nurse (name):**

Start consultation time:	End time:

Examinations done

- ☐ Blood Pressure
- ☐ Heart rate
- ☐ Oxygen saturation
- ☐ Weight
- ☐ Height
- ☐ Smoking status
- ☐ Alcohol consumption

Calculations done

- ☐ BMI
- ☐ ACR

Was the patient referred to a service (e.g. COACH)?  Yes / No         Service Name: ……

Was counselling provided during the interaction?	Yes / No

Comments

**Step 2: Doctor Consultation**

Name of Dr:	       Training level: Res / Reg / Consultant             Gen med / Cardio

Time spent reviewing referral prior to consult:

Start consultation time:	End time:

Risk factors discussed & number of times discussed

- ☐ Smoking    …………… *times*
- ☐ Alcohol consumption    …………… *times*
- ☐ Exercise   …………… *times*
- ☐ Diet   …………… *times*
- ☐ Other …………….

Was the Absolute Cardiovascular Risk Score discussed during the consult?	Yes / No

Comments

Examinations conducted during consultation

- ☐ Cardiovascular examination
- ☐ Respiratory examination
- ☐ Other, please specify  …………….

Comments

**Step 3: Follow up triage decisions**

Patient commenced on medication	**Yes** / No 	If Yes, name ……Statins…(pt was very apprehensive but was counselled on the side effects extensively…….

Were the patient referred for further investigations	**Yes** / No

If yes, specify which type

- ☐ Stress Echo
- ☐ Exercise stress test : *initially considered*
- ☐ Cardiac CT
- ☐ MIBI
- ☐ Other, please specify:  ……………………………

Was the patient counselled on the test (how it works, location, price, waiting time?)

How will the patient be contacted about the follow-up results?

- ☐ Contacted from RACPC
- ☐ Recommended to organise appointment from GP
- ☐ Informed at the site when the patient completes testing

**Step 4: Clinical Note Taking**

Which components of the discharge summary was completed by the doctor?

- ☐ Situation
- ☐ Observation
- ☐ Background (Smoking status)
- X Lifestyle assessment
- ☐ Drug Allergies
- ☐ Assessment
- Clinical symptoms
- ☐ Problems/ diagnosis
- ☐ Prescriptions
- ☐ Vaccination log
- ☐ Procedures
- ☐ Investigations
- ☐ Recommendations

Comments

**APPENDIX B. RACPAC HEALTHCARE PRACTITIONER / CLINIC STAFF INTERVIEW QUESTIONS**

**What are the perspectives of RACPAC doctors, nurses and staff of the level of service which the clinic offers to patients?**

The RACPAC was established to provide a high standard of care to patients. The perceptions of those working within the service will be fundamental to determining whether the clinic succeeds in this goal. Health practitioner and staff confidence in the level of care offered by the RACPAC service will to be determined by questions such as these:

Interview prompts:

- Prior to RACPAC’s establishment how were patients that presented with chest pain (either through the ED or GP referrals) handled by the Royal Hobart hospital?
- What are the most noticeable aspects of the RACPAC service which are different to usual care?
- How confident are you that the RACPAC offers a high level of care to its patients? In what way does it do this (what protocols are in place)?
- In your opinion is the well-being of patients better managed through the RACPAC or the hospital’s usual care.
- How has the delivery of RACPAC changed in response to the COVID-19 pandemic?
- Have any of the changes to the delivery of RACPAC in response to the COVID-19 pandemic impacted the quality of patient care or patient experience?

**Do RACPAC doctors and staff believe that the Clinic’s processes are sufficiently** **streamlined for an effective service?**

One specific purpose of the RACPAC is to provide a more efficient healthcare service for its patients, one that is a notable improvement on the long wait-times and inefficient follow-up at the Royal Hobart Hospital prior to the Clinic’s establishment. While research has shown the clinic to indeed be more efficient in handling chest pain patients [1], it is important to understand how the RACPAC ensures this efficiency within its daily processes, whether staff think efficiency is maximised, and whether any improvements can be made.

Interview prompts:

- Please explain how the clinic operates on a daily/weekly basis.
- What is required to for the Clinic to run? How many staff are required?
- Do you believe the efficiency of the RACPAC is maximised? Why/why not?
- Is there anything unique about this service that you think makes it more streamlined than other models of care?
- Are there any improvements that can be made to the current processes?

**What are the perceptions of RACPAC management and staff regarding how the Clinic** **is run?**

For the purpose of sustainability, it is important to understand in more detail how the Clinic is run. This interview theme will provide a clear understanding of how the RACPAC is managed. The following questions will also determine if there are issues regarding the RACPAC’s management style, resourcing and/or communication within the Clinic.

Interview prompts:

- How would you describe the Clinic’s management style?
- Is the Clinic’s resourcing sufficient for it to run effectively? Resources could refer to staffing, time per patient, consultation rooms, etc.
- If doctors and staff have suggestions or concerns about how the RACPAC is being run, is there a channel of communication available to voice their opinions? Please provide examples if possible.
- Does the Clinic’s setting within Royal Hobart Hospital influence it’s functioning?
- RACPAC autonomy - to what extent is the clinic influenced (if at all) by the broader management of the Royal Hobart Hospital?

**Positive and negative attributes**

This interview theme will attempt to isolate perceptions about the most important element/s of the RACPAC to maintain and which to discard.

Interview prompts:

- Please identify the most important positive and negative attribute of the clinic. How would you suggests maintaining the positive attribute and improving the negative attribute?

**APPENDIX C. FRAMEWORK ANALYSIS CODEBOOK**

| **Table. Context, mechanisms, and outcomes obtained from the thematic analysis of the document review to support in-depth interviews with RACPC clinicians [1-15].** | | |
| --- | --- | --- |
| **Context** | **Mechanisms** | **Outcomes** |
| Distal (Macro-level) Context | Management/Provider level | Long-term outcomes |
| - Stakeholder collaboration | - Perceived social need and support | - Healthier Tasmanians - Intervention ‘standardisation’ - Staff retention, acceptability and satisfaction |
| - Leadership and higher-level support | - Motivation - Positive peer dynamics |  |
| - Monitoring, evaluation and research | - Continued quality improvement and safety |  |
| Organisational (Meso-level) Context | Social mechanisms | Immediate outcomes |
| - Implementation methodology | - Patient-centred, patient-empowered - Risk-stratified (patient factors) - Active involvement in RACPC care (patients/providers) | - Improvement patient self-management and treatment compliance |
| - Staffing components (multidisciplinary, number, dynamics, education and training) | - Composition: Cardiologists, nurses, medical officers, administrative support - Collaborative, team-based approach - Mutual learning: recognition and confidence in teams’ competencies - Internalisation of collective leadership and decision-making | - Reduced operational staff workload (shared) - Decongestion of clinic and wait times - Staff retention, acceptability and satisfaction - Fostered culture of learning, collaboration and continuous quality improvement |
| - Overarching hierarchical pressure (Health Service oversight) | - Social/peer and/or organisational support - Commitment to service identity and values - Recognition of service’s place in outpatient cardiac care | - Supported organisational efficiency and effectiveness (decongestion of clinic) - Improved patient access to care |
| Local (Micro-level) Context | Provider level | Immediate outcomes |
| - Clinical environment | - Conducive environment - Continuity of care | - Decreased staff satisfaction and productivity (decreased workload for operational staff) - Decreased patient satisfaction, acceptability and opportunity cost - Impacted organisational inefficiencies to patient care |
| - Administrative burdens |  |  |
| - RACPC service champions | - Trust, communication and support | - Sustained service need and delivery |
